# Contact investigation of tuberculosis in Brunei Darussalam: Evaluation and risk factor analysis

**DOI:** 10.1101/2022.02.04.22270445

**Authors:** Liling Chaw, Rafizah Abdul Hamid, Kai Shing Koh, Kyaw Thu

## Abstract

**Introduction:** We evaluated the yield of tuberculosis (TB) contact investigation in Brunei Darussalam, and identified the associated factors for latent TB infection (LTBI) diagnosis, as well as for initiating and completing LTBI treatment.

**Methods:** Data was extracted and digitalised for all close contacts of pulmonary TB cases at the National TB Coordinating Centre from January 2009 to December 2018. Generalising estimating equations (GEE) logistic regression models were used to determine the associated factors. Manual matching against electronic health records system was done to identify contacts who had progressed to active TB disease.

**Results:** Among 10,537 contacts, 9.9% (n= 1047) were diagnosed as LTBI, out of which 43.0% (n= 450) initiated LTBI treatment. Among those who initiated, 74.0% (n= 333) completed LTBI treatment. Contact factors associated with LTBI diagnosis include being male (adjusted Odds Ratio (aOR)= 1.18 [95%CI: 1.03,1.34]), local (aOR= 0.70 [95%CI: 0.56,0.88]), and a household contact (aOR= 1.59 [95%CI: 1.26,1.99]). Contacts of index cases who were <60 years old and diagnosed as smear positive PTB (aOR= 1.62 [95%CI: 1.19,2.20]) had higher odds of being diagnosed with LTBI. Local LTBI cases had higher odds of initiating LTBI treatment (aOR= 1.86 [95%CI: 1.26,2.73]). Also, LTBI cases detected from local (aOR= 2.32 [95%CI: 1.08,4.97]) and smear positive PTB index cases (aOR= 2.23 [95%CI: 1.09,4.55]) had higher odds of completing LTBI treatment. Among 1047 LTBI cases, 5 (0.5%) had progressed to active PTB within 1-8 years post-LTBI diagnosis.

**Discussion:** LTBI burden is disproportionately high towards foreign nationals, with higher odds of LTBI diagnosis but lower odds of treatment initiation. Determining the reasons of not initiating LTBI treatment will be useful to help improve LTBI treatment uptake. Establishing digital databases and building TB laboratory capacity for molecular typing would be useful to determine the contribution of LTBI or reactivation towards TB incidence in Brunei.

**Key messages:** *What is already known on this topic:*

- Brunei is an intermediate TB burden country with stagnating TB incidence since 2004, despite having an established national TB programme.
- We undertook this study to evaluate the contact investigation process and also to understand the contribution of LTBI towards TB incidence in the country.

*What this study adds:*

- Among contacts of PTB index cases identified between 2009 and 2018, the proportions of LTBI diagnosis, treatment initiation and treatment completion were 9.9%, 43.0% and 74.0%, respectively.
- The associated factors of LTBI diagnosis include being, male, foreign and a household contact. Local cases had higher odds of initiating LTBI treatment when compared to foreign nationals.

*How this study might affect research, practice or policy:*

- Strengthening LTBI screening and treatment uptake among foreign nationals could be an important step towards reducing TB incidence in Brunei.

## Introduction

Tuberculosis (TB) still remains a global public health concern, with an estimated 10 million new cases and about 1.2 million deaths in 2019 [1] during the pre-COVID pandemic period. Contact investigation is an active case-finding approach that involves the systematic evaluation of the contacts of identified TB patients to identify active TB disease or latent TB infection (LTBI) [2]. The rationale is that close contacts are at high risk of TB infection when compared to the general population, and that those exposed to infectious TB cases tend to progress to active disease within 1-2 years post-infection [3]. Contacts who tested positive for LTBI are being offered preventive LTBI treatment, with an aim to eliminate the infection before progressing to TB disease.

Brunei Darussalam (pop. 453,600 [4]) is a small Southeast Asian country with a TB incidence of 57 per 100,000 population in 2017 [5]. Since 2000, the country has implemented the National TB Control Programme (NTP), with an end goal of eliminating TB by 2050 [6]. Following the WHO Stop TB strategy [7], contact investigation plays a key role in TB control in Brunei, and is indicated for contacts of TB cases with pulmonary or laryngeal disease.

Contact investigations have been evaluated particularly in low TB burden countries like the Netherlands [8] and England [9], where the focus is to detect and treat as many LTBI cases as possible. Despite being routinely conducted, the yield for TB contact investigation has never been evaluated in Brunei; one reason being that records were entirely hardcopy-based. Also, understanding both the extent and risk factors of developing LTBI among contacts may also help us to uncover the causes of stagnating TB incidence in the country since 2004 [6]. Hence, this study aims to: (1) evaluate the contact investigation process in Brunei, using established indicators, (2) investigate the associated factors of being diagnosed with LTBI among close contacts of PTB cases, (3) identify the determinants of LTBI treatment initiation & completion, and lastly, (4) describe the LTBI cases who later progressed to active TB disease.

The study findings could be useful to assess the outcomes of the current contact investigation process, and identify ways for further improvements. Also analysing such dataset provides an opportunity to determine risk factors of LTBI in enclosed settings.

## Methods

### Study design and the contact investigation process

We conducted a retrospective cohort study covering all registered PTB patients and their close contacts in Brunei Darussalam from January 2009 to December 2018. Data was collected from the National Tuberculosis Coordinating Centre (NTCC), a centre within the country’s Ministry of Health that was established to implement, monitor, coordinate, and evaluate TB prevention and control programs. Being a notifiable disease under Brunei’s Infectious Disease Act [10], it is required to report all suspected TB patients identified in either public or private healthcare settings to the Ministry of Health. Hence, all suspected TB patients in the country (including those who visited private clinics and hospitals) are referred to NTCC or any of the directly observed treatment, short-course (DOTS) centres located in all 4 districts for diagnosis, treatment, and follow-up. NTCC and all district DOTS centres also conduct active contact tracing for all diagnosed and suspected active PTB cases, using the ‘stone in the pond’ principle starting with higher-risk contacts and expanded further according to risk assessment. Source case investigation was also conducted for child TB cases under 5 years old. During a routine contact investigation, the period of inquiry for contact exposure starts from 3 months prior to either the diagnosis date or date of cough onset (if known). This period would be extended if the index case was considered highly infectious. High-risk close contacts included the immediate household members and others who have shared accommodation with the index case. Contacts at work, leisure or other settings were also screened, based on risk assessment conducted during the contact investigation.

Tuberculin Skin Test (TST) was the main test used to determine LTBI status and was used to screen all close contacts identified during the contact investigation [6]. During periods of purified protein derivative (PPD) shortage (between 2015 and 2018), priority was given to contacts of smear positive PTB and child TB cases. TST positive contacts who were symptomatic were assessed by routine investigation (chest X-ray and 3 consecutive sputum collections) to exclude active TB disease. When there was no evidence of active TB in TST positive contacts, they were diagnosed as LTBI and were offered LTBI treatment (Isoniazid daily for 6 months). They were reviewed by LTBI clinic every 6-8 weeks and supplied medications on a monthly basis. This treatment was provided free of charge for both local and foreign nationals, however, it was not mandatory.

### Data collection

Two separate datasets were compiled. First, epidemiological and clinical data from all PTB index cases were extracted from the NTCC database. Data collected include socio-demographics (age, gender, nationality, district of residence), year of diagnosis, and type of PTB. The latter (type of PTB) was divided into 3 categories: the first 2 based on sputum smear microscopy results (smear positive PTB and smear negative PTB), and other PTB which consisted of patients who were unable to provide adequate sputum samples and were instead diagnosed through other methods (such as radiological imaging, clinical judgement, gastric lavage and tissue biopsy). Second, data all close contacts were digitalised from available hardcopy records of contact investigations. The collected data include socio-demographics (age, gender, nationality, district of residence), index case number, type of contact (household, workplace, school or others), dates and results of the first and/or second TST test. A household contact was defined as one who had shared accommodation with the index case (hence having the same residential address). For contacts diagnosed as LTBI, further data was collected as follows: assigned LTBI case number, LTBI diagnosis date, LTBI treatment start and end dates, and reasons for not completing treatment.

All recent contacts were diagnosed as LTBI when they have a TST reading of ≥10mm (prior to April 2013) or ≥5mm (after April 2013), following the cut-off values before and after the revision of the national TB guidelines [6]. The revised cut-off value or ≥5mm was based on recommendations by the United States Centers of Disease Control and Prevention, with the aim of incorporating a risk categorisation component to the LTBI definition [12, 13]. A TST test was considered to be given if there were records of taking a TST test, and/or if a TST test result was recorded. Contacts with LTBI were considered as started and/or completed their LTBI treatment if a treatment start and/or end date was recorded.

To identify contacts with LTBI who eventually progressed to active TB, manual matching using the name and/or national identity card number was performed by a clinician (and co-author) against the Brunei Health Information Management System database (Bru-HIMS). Bru-HIMS is a comprehensive electronic health record system and was used in all government hospitals and clinics since 2013 [14]. If a match was found, information on the date of PTB diagnosis, type of PTB, and family history of TB was collected from Bru-HIMS.

### Statistical analysis

This study has 3 outcomes of interest: 1) the proportion of contacts with LTBI diagnosis (the LTBI yield), 2) the proportion of LTBI cases who initiated LTBI treatment, and 3) the proportion of contacts who completed LTBI treatment. First, each proportion was calculated using the following as the denominators: total number of contacts tested for LTBI (for proportion 1), total number of contacts diagnosed with LTBI (for proportion 2), and total number of contacts who were diagnosed with LTBI and also initiated LTBI treatment (for proportion 3).

Next, the demographic and clinical determinants for both contacts and index were initially assessed using independent t, chi-squared or Fisher’s exact tests, wherever appropriate. As multiple contacts were traced from the same index case, we used generalising estimating equations (GEE) logistic regression models to identify associated factors while accounting for clustered data, using an exchangeable correlation matrix. Variables with univariate p-values of < 0.2 were included into the multivariable analysis. Quasi Information Criterion (QIC) values were used to evaluate models, with smaller QIC values indicating better model fit. Variables were subsequently removed from multivariable models if they did not have an independent association with the outcome, its inclusion increased the QIC value, and/or their exclusion did not affect the estimates of other variables. Age and gender (of both contact and index cases) were kept in multivariable models a priori. All analyses were done using R (ver. 4.1.0) [15] and the geepack package [16]. A p-value ≤ 0.05 was considered as statistically significant.

As GEE analysis requires all missing data to be removed, the total number of contacts were different for all such analyses in the study. To avoid reporting bias, we opt to report the overall proportions of LTBI diagnosis, LTBI treatment initiation and completion using the whole dataset. Ethical approval was given by the Medical and Health Research and Ethics Committee, Ministry of Health, Brunei (Ref: MHREC/UBD/2019/2).

## Results

A total of 11,749 contacts from 1,088 PTB index cases were identified between January 2009 and December 2018. Contacts whose index cases who were later diagnosed as EPTB (261, 2.2%), with no recorded TST results (392, 3.3%), and from one outlier case (559, 4.8%) were excluded from the study (Figure 1). After these exclusions, our final dataset consisted of 10,537 contacts (89.7%) from 1,048 PTB index cases.

**Figure 1.**
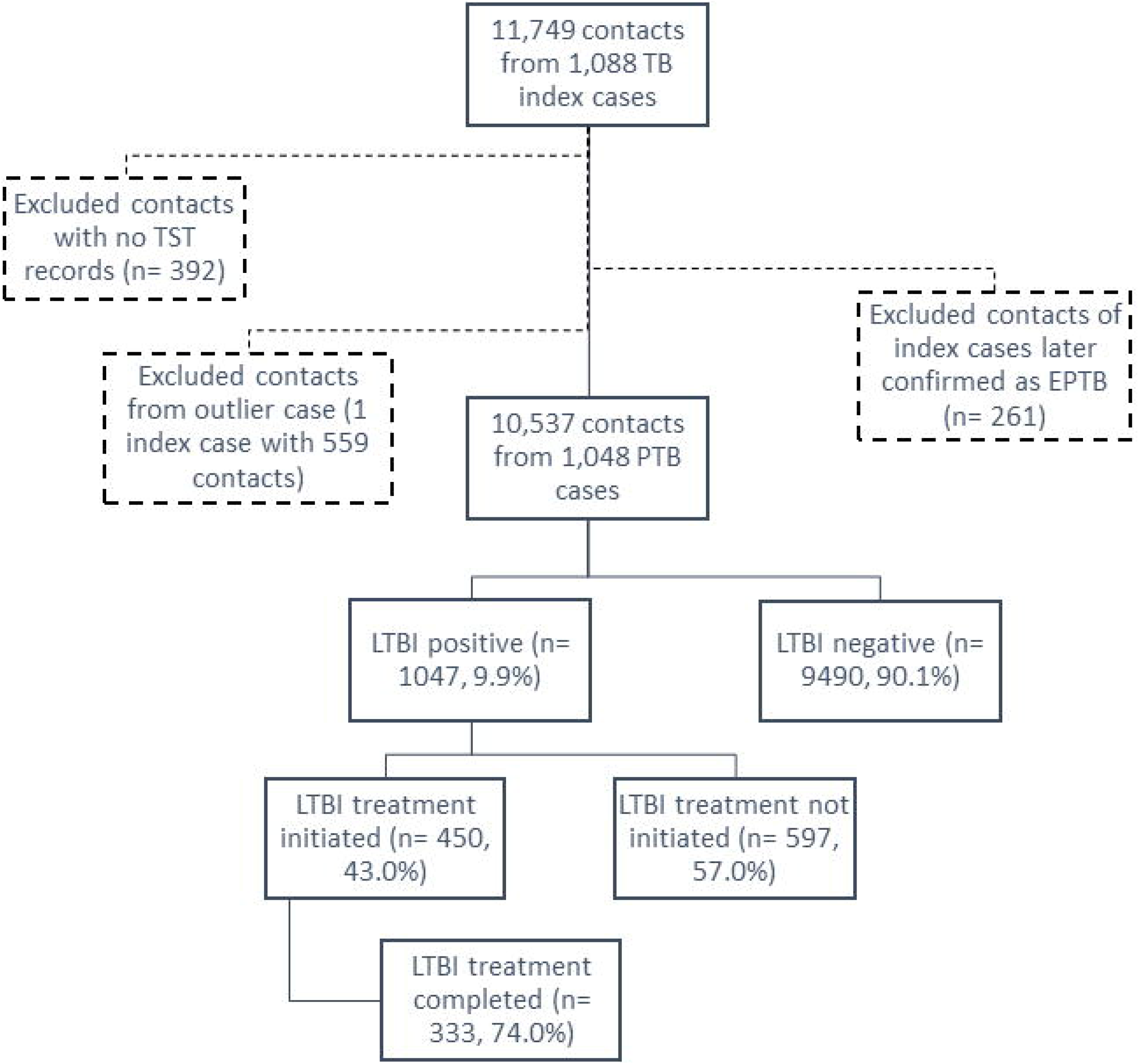
Study flowchart

### Characteristics of index PTB cases and overall contacts

The median age (IQR) of the index PTB cases was 47.0 years (32 - 62), ranging between 1 to 95 years (Table 1). The majority were male (671, 64.0%). Their median (IQR) number of contacts was 6.0 (3 - 12), ranging between 1 to 101 individuals. The majority were diagnosed as smear positive PTB (693, 66.1%).

**Table 1.**
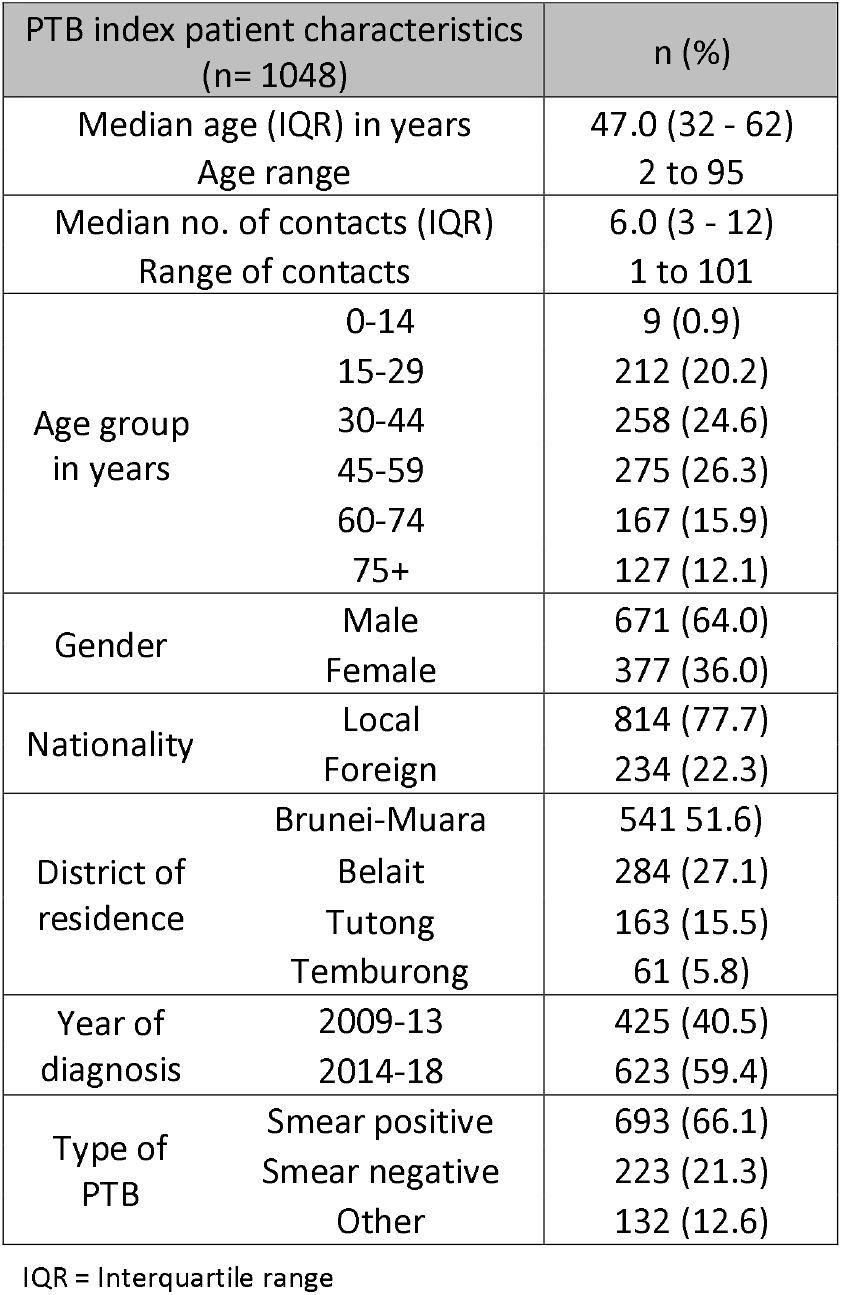
Characteristics of the index PTB patients registered from 2009 to 2018.

| PTB index patient characteristics<br>(n= 1048) |  | n (%) |
| --- | --- | --- |
| Median age (IQR) in years |  | 47.0 (32 - 62) |
| Age range |  | 2 to 95 |
| Median no. of contacts (IQR) |  | 6.0 (3 - 12) |
| Range of contacts |  | 1 to 101 |
| Age group<br>in years | 0-14 | 9 (0.9) |
|  | 15-29 | 212 (20.2) |
|  | 30-44 | 258 (24.6) |
|  | 45-59 | 275 (26.3) |
|  | 60-74 | 167 (15.9) |
|  | 75+ | 127 (12.1) |
| Gender | Male | 671 (64.0) |
|  | Female | 377 (36.0) |
| Nationality | Local | 814 (77.7) |
|  | Foreign | 234 (22.3) |
| District of<br>residence | Brunei-Muara | 541 (51.6) |
|  | Belait | 284 (27.1) |
|  | Tutong | 163 (15.5) |
|  | Temburong | 61 (5.8) |
| Year of<br>diagnosis | 2009-13 | 425 (40.5) |
|  | 2014-18 | 623 (59.4) |
| Type of<br>PTB | Smear positive | 693 (66.1) |
|  | Smear negative | 223 (21.3) |
|  | Other | 132 (12.6) |

IQR = Interquartile range

The median age (IQR) of the contacts was 31.0 years (19 - 42), ranging between 6 months to 98 years (Table 2). The majority were local residents (8597, 81.6%). About half of them were males (5474, 52.0%), residing in Brunei-Muara district (5007, 47.5%) and classified as household contacts (5192, 49.3%).

**Table 2.**
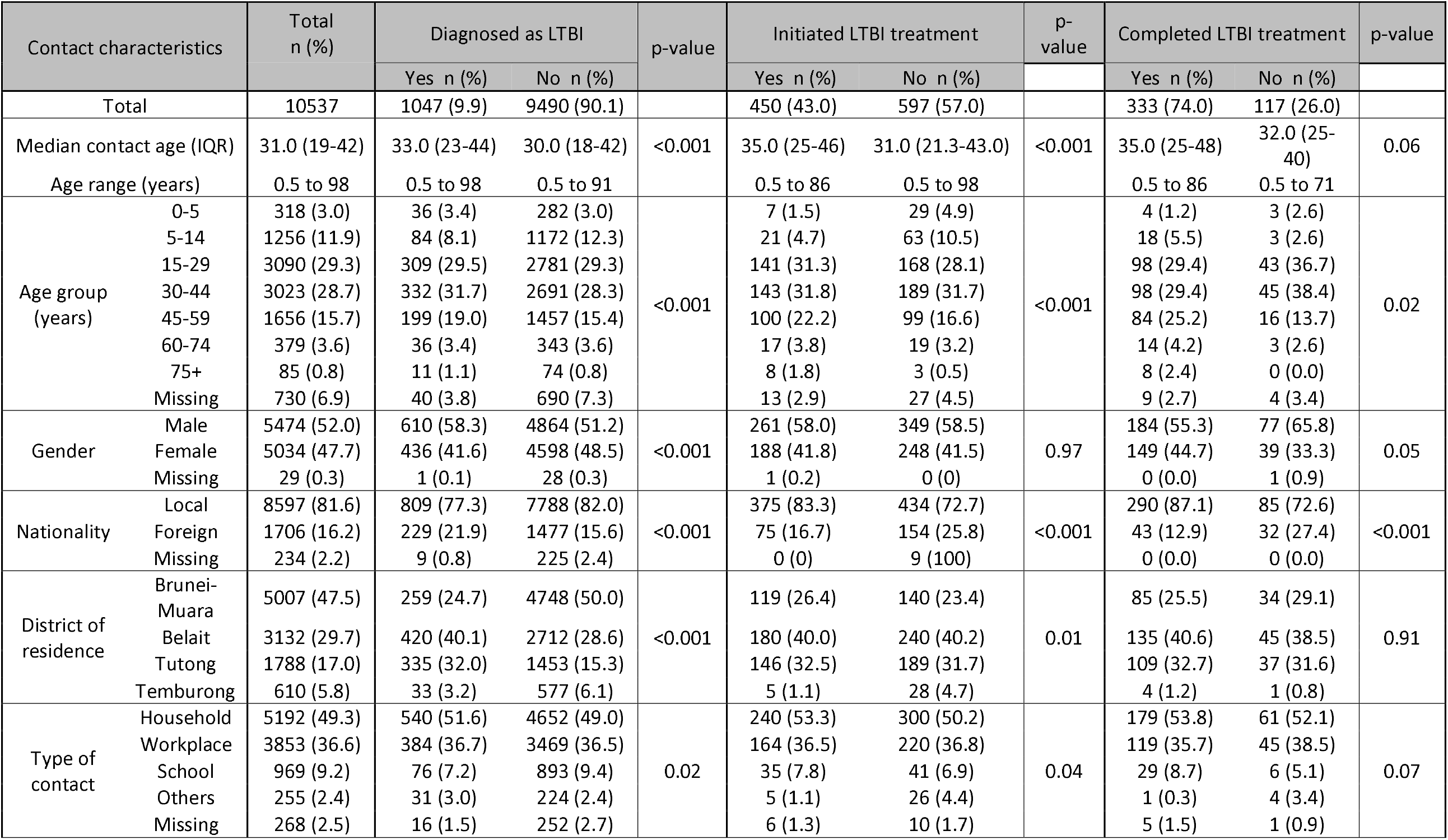
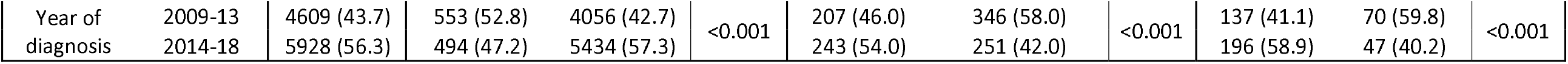
Characteristics of close contacts registered from 2009 to 2018.

### Associated factors of LTBI diagnosis among contacts

Among all contacts, 1047 (9.9%) were diagnosed as LTBI. Significant differences were observed for all assessed characteristics between the LTBI and non-LTBI groups (Table 2). Multivariable analysis (Table 3) showed that male contacts had higher odds of being diagnosed with LTBI (adjusted Odds Ratio (aOR)= 1.18 [95% Confidence Interval (95% CI): 1.03, 1.34]).

**Table 3.**
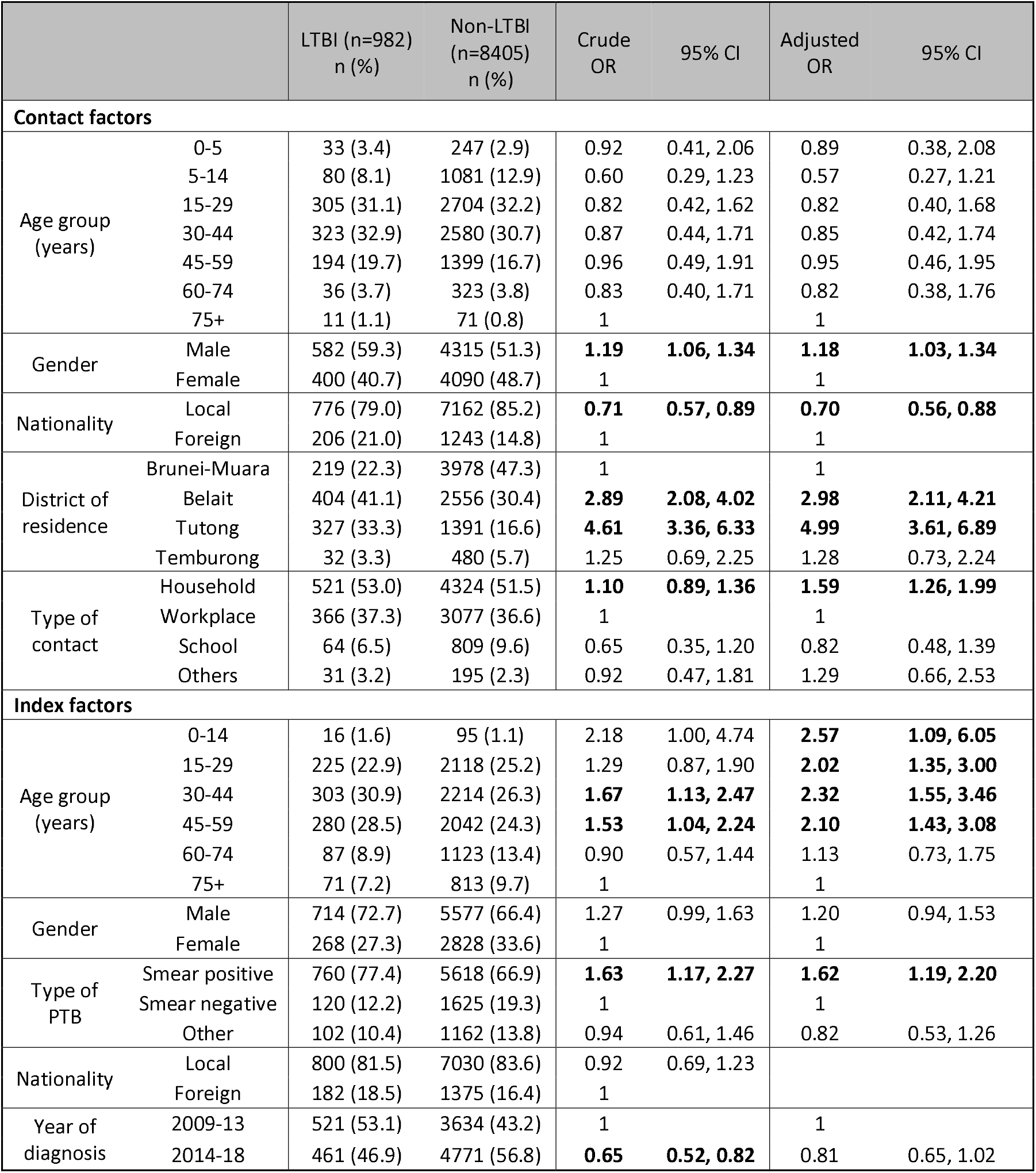
Factors associated with LTBI diagnosis among close contacts from 2009 to 2018 [n= 9387 out of 10537 (89.1%)].

Other contact factors associated with LTBI diagnosis are: local residents when compared to foreign (aOR= 0.70 [95% CI: 0.56, 0.88]), contacts who resided in Belait (aOR= 2.98 [95% CI: 2.11, 4.21]) or Tutong (aOR= 4.99 [95% CI: 3.61, 6.89]) when compared to Brunei-Muara district, and household contacts of the index case when compared to workplace contacts (aOR= 1.59 [95% CI: 1.26, 1.99]).

Contacts of index cases aged < 60 years had more than twice the odds of being diagnosed with LTBI, when compared to those ≥ 75 years, Table 3). Contacts of index cases who were diagnosed as smear positive PTB also had higher odds of being diagnosed with LTBI, when compared to smear negative (aOR= 1.62 [95% CI: 1.19,2.20]).

### LTBI treatment initiation among contacts diagnosed with LTBI

Among LTBI cases (n= 1047), 450 (43.0%) initiated LTBI treatment. Significant differences were observed between both groups, except for contact’s gender (Table 2). Multivariable analysis (Table 4) revealed that contacts < 15 years old had at least 78% lower odds of initiating LTBI treatment when compared to those ≥ 75 years. Also, local LTBI cases had higher odds of initiating LTBI treatment, when compared to foreign residents (aOR= 1.86 [95% CI: 1.26, 2.73]). When compared to Brunei-Muara, those residing in Temburong district had lower odds of initiating LTBI treatment (aOR= 0.20 [95% CI: 0.07, 0.61]).

**Table 4.** Factors associated with LTBI treatment initiation among contacts diagnosed with LTBI from 2009 to 2018 [n= 982 out of 1047 (93.8%)].

|  |  | LTBI treatment initiated<br>(n=430)<br>n (%) | LTBI treatment not initiated<br>(n=552)<br>n (%) | Crude OR | 95% CI | Adjusted OR | 95% CI |
| --- | --- | --- | --- | --- | --- | --- | --- |
| **Contact factors** |  |  |  |  |  |  |  |
| Age group (years) | 0-5 | 7 (1.6) | 26 (4.7) | 0.29 | 0.07, 1.17 | **0.17** | **0.04, 0.81** |
|  | 5-14 | 21 (4.9) | 59 (10.7) | 0.30 | 0.09, 1.02 | **0.22** | **0.05, 0.87** |
|  | 15-29 | 141 (32.8) | 164 (29.7) | 0.76 | 0.24, 2.48 | 0.67 | 0.18, 2.54 |
|  | 30-44 | 140 (32.5) | 183 (33.2) | 0.64 | 0.20, 2.06 | 0.58 | 0.15, 2.19 |
|  | 45-59 | 96 (22.3) | 98 (17.8) | 0.81 | 0.25, 2.68 | 0.77 | 0.20, 3.00 |
|  | 60-74 | 17 (4.0) | 19 (3.4) | 0.74 | 0.20, 2.75 | 0.49 | 0.11, 2.15 |
|  | 75+ | 8 (1.9) | 3 (0.5) | 1 |  | 1 |  |
| Gender | Male | 254 (59.1) | 328 (59.4) | 1.08 | 0.84, 1.40 | 1.18 | 0.90, 1.54 |
|  | Female | 176 (40.9) | 224 (40.6) | 1 |  | 1 |  |
| Nationality | Local | 361 (84.0) | 415 (75.2) | **1.57** | **1.11, 2.23** | **1.86** | **1.26, 2.73** |
|  | Foreign | 69 (16.0) | 137 (24.8) | 1 |  | 1 |  |
| District of residence | Brunei-Muara | 107 (24.9) | 112 (20.3) | 1 |  | 1 |  |
|  | Belait | 176 (40.9) | 228 (41.3) | 0.80 | 0.53, 1.20 | 0.83 | 0.54, 1.28 |
|  | Tutong | 142 (33.0) | 185 (33.5) | 0.75 | 0.49, 1.15 | 0.72 | 0.46, 1.13 |
|  | Temburong | 5 (1.2) | 27 (4.9) | **0.25** | **0.09, 0.73** | **0.20** | **0.07, 0.61** |
| Type of contact | Household | 235 (54.7) | 286 (51.8) | 1.05 | 0.72, 1.54 | 1.15 | 0.74, 1.79 |
|  | Workplace | 159 (37.0) | 207 (37.5) | 1 |  | 1 |  |
|  | School | 31 (7.2) | 33 (6.0) | 1.13 | 0.40, 3.26 | 1.23 | 0.43, 3.54 |
|  | Others | 5 (1.1) | 26 (4.7) | 0.44 | 0.14, 1.41 | 0.38 | 0.11, 1.26 |
| **Index factors** |  |  |  |  |  |  |  |
| Age group (years) | 0-14 | 9 (2.1) | 7 (1.3) | 0.93 | 0.16, 5.32 | 1.07 | 0.21, 5.43 |
|  | 15-29 | 98 (22.8) | 127 (23.0) | 0.82 | 0.45, 1.46 | 0.89 | 0.48, 1.65 |
|  | 30-44 | 131 (30.5) | 172 (31.1) | 0.85 | 0.48, 1.52 | 1.02 | 0.56, 1.86 |
|  | 45-59 | 110 (25.5) | 170 (30.8) | 0.72 | 0.40, 1.29 | 0.72 | 0.39, 1.34 |
|  | 60-74 | 48 (11.2) | 39 (7.1) | 1.10 | 0.55, 2.18 | 1.46 | 0.72, 2.94 |
|  | 75+ | 34 (7.9) | 37 (6.7) | 1 |  | 1 |  |
| Gender | Male | 304 (70.7) | 410 (74.3) | 0.91 | 0.64, 1.31 | 0.99 | 0.67, 1.47 |
|  | Female | 126 (29.3) | 142 (25.7) | 1 |  | 1 |  |
| Type of PTB | Smear positive | 334 (77.6) | 426 (77.2) | 1.15 | 0.71, 1.85 |  |  |
|  | Smear negative | 48 (11.2) | 72 (13.0) | 1 |  |  |  |
|  | Other | 48 (11.2) | 54 (9.8) | 1.18 | 0.61, 2.25 |  |  |
| Nationality | Local | 362 (84.2) | 438 (79.3) | 1.36 | 0.89, 2.06 |  |  |
|  | Foreign | 68 (15.8) | 114 (20.7) | 1 |  |  |  |
| Year of diagnosis | 2009-13 | 198 (46.0) | 323 (58.5) | 1 |  | 1 |  |
|  | 2014-18 | 232 (54.0) | 229 (41.5) | **1.43** | **1.03, 1.99** | 1.39 | 0.98, 1.98 |

### LTBI treatment completion among contacts who initiated treatment

Among those who initiated LTBI treatment (n= 450), 333 (74.0%) completed it. Significant differences for contact’s age group, nationality and the year of contact investigation were observed between both groups (Table 2). Multivariable analysis (Table 5) showed that contacts from index cases of < 15 years old had lower odds of completing LTBI treatment when compared to those ≥ 75 years (aOR= 0.06 [95% CI: 0.02, 0.22]). Also, LTBI cases detected from local (aOR= 2.32 [95% CI: 1.08, 4.97]) and smear positive PTB index cases (aOR= 2.23 [95% CI: 1.09, 4.55]) had higher odds of completing LTBI treatment. Lastly, the latter half of the study period (2014-2018) was positively associated with LTBI treatment completion (aOR= 1.95 [95% CI: 1.10, 3.46]).

**Table 5.** Factors associated with LTBI treatment completion among close contacts who initiated LTBI treatment from 2009 to 2018 [n = 430 out of 450 (95.6%)].

|  |  | LTBI treatment completed<br>(n=319)<br>n (%) | LTBI treatment not completed<br>(n=111)<br>n (%) | Crude OR | 95% CI | Adjusted OR | 95% CI |
| --- | --- | --- | --- | --- | --- | --- | --- |
| **Contact factors** |  |  |  |  |  |  |  |
| Age group (years) | 0-14 | 22 (6.9) | 6 (5.4) | 0.47 | 0.09, 2.45 | 0.34 | 0.04, 2.82 |
|  | 15-29 | 98 (30.7) | 43 (38.7) | 0.30 | 0.07, 1.23 | 0.29 | 0.05, 1.84 |
|  | 30-44 | 97 (30.4) | 43 (38.7) | 0.27 | 0.07, 1.05 | 0.24 | 0.04, 1.44 |
|  | 45-59 | 80 (25.1) | 16 (14.5) | 0.49 | 0.12, 2.01 | 0.53 | 0.08, 3.41 |
|  | 60+ | 22 (6.9) | 3 (2.7) | 1 |  | 1 |  |
| Gender | Male | 180 (56.4) | 74 (66.7) | 0.73 | 0.48, 1.13 | 0.88 | 0.52, 1.50 |
|  | Female | 139 (43.6) | 37 (33.3) | 1 |  | 1 |  |
| Nationality | Local | 279 (87.5) | 82 (73.9) | **2.50** | **1.32, 4.74** |  |  |
|  | Foreign | 40 (12.5) | 29 (26.1) | 1 |  |  |  |
| District of residence | Brunei-Muara | 77 (24.1) | 30 (27.0) | 1 |  |  |  |
|  | Belait | 131 (41.1) | 45 (40.6) | 1.23 | 0.68, 2.22 |  |  |
|  | Tutong | 107 (33.5) | 35 (31.5) | 1.32 | 0.73, 2.40 |  |  |
|  | Temburong | 4 (1.3) | 1 (0.9) | 1.66 | 0.18, 15.51 |  |  |
| Type of contact | Household | 178 (55.8) | 57 (51.4) | 1.30 | 0.73, 2.31 |  |  |
|  | Workplace | 115 (36.1) | 44 (39.6) | 1 |  |  |  |
|  | School | 25 (7.8) | 6 (5.4) | 1.81 | 0.53, 6.18 |  |  |
|  | Others | 1 (0.3) | 4 (3.6) | 0.30 | 0.03, 3.02 |  |  |
| **Index factors** |  |  |  |  |  |  |  |
| Age group (years) | 0-14 | 3 (1.0) | 6 (5.4) | **0.05** | **0.01, 0.17** | **0.06** | **0.02, 0.22** |
|  | 15-29 | 68 (21.3) | 30 (27.0) | **0.20** | **0.05, 0.71** | 0.30 | 0.08, 1.19 |
|  | 30-44 | 98 (30.7) | 33 (29.8) | 0.28 | 0.08, 1.02 | 0.45 | 0.11, 1.78 |
|  | 45-59 | 83 (26.0) | 27 (24.3) | 0.32 | 0.09, 1.19 | 0.37 | 0.09, 1.49 |
|  | 60-74 | 36 (11.3) | 12 (10.8) | 0.34 | 0.09, 1.33 | 0.35 | 0.09, 1.45 |
|  | 75+ | 31 (9.7) | 3 (2.7) | 1 |  | 1 |  |
| Gender | Male | 216 (67.7) | 88 (79.3) | 0.65 | 0.36, 1.19 | 0.75 | 0.41, 1.38 |
|  | Female | 103 (32.3) | 23 (20.7) | 1 |  | 1 |  |
| Type of PTB | Smear positive | 262 (82.1) | 72 (64.9) | 1.82 | 0.90, 3.66 | **2.23** | **1.09, 4.55** |
|  | Smear negative | 31 (9.7) | 17 (15.3) | 1 |  | 1 |  |
|  | Other | 26 (8.2) | 22 (19.8) | 0.53 | 0.20, 1.37 | 0.73 | 0.25, 2.13 |
| Nationality | Local | 282 (88.4) | 80 (72.1) | **2.88** | **1.51, 5.50** | **2.32** | **1.08, 4.97** |
|  | Foreign | 37 (11.6) | 31 (27.9) | 1 |  | 1 |  |
| Year of | 2009-13 | 131 (41.1) | 67 (60.4) | 1 |  | 1 |  |
| diagnosis | 2014-18 | 188 (58.9) | 44 (39.6) | **1.70** | **1.01, 2.86** | **1.95** | **1.10, 3.46** |

### LTBI cases who progressed to active TB disease

Among 1047 LTBI cases, 5 (0.5%) eventually progressed to active PTB within 1 to 8 years post-LTBI diagnosis. Among these 5 cases, 1 is a foreign national and 2 had history of TB infection among their household members. All 5 have developed smear-positive PTB (out of which only 2 developed the disease within the first 2 years of LTBI diagnosis). All except one had previously completed LTBI treatment.

## Discussion

During the 10-year study period, we found that the prevalence of LTBI among all screened contacts was 9.9%. This finding is low when compared to that reported from a systematic review for high-income settings (28.1%), but comparable to that from low TB-burden countries of the Netherlands (11%) and among child contacts in England (10%) [2, 8, 9]. We found that contact factors associated with increased odds of LTBI diagnosis include being foreign, male, and a household contact of index PTB cases who were < 60 years old and diagnosed with smear positive PTB. Foreign residents [17, 18], household contacts [18–20], and male gender [20, 21] have previously been shown to be at increased risk of LTBI diagnosis. Also, contacts of smear positive PTB cases have been previously reported as having higher risk of LTBI diagnosis [8]; cough aerosol experiments have showed smear positive PTB cases as a risk factor of infectiousness [22].

Despite foreign nationals constituting 16.2% of the total contact population in our study, their percentage of LTBI diagnosis (13.4%) was higher when compared to that of the locals (9.4%). This indicates that foreign nationals were highly and disproportionately affected by LTBI. Yet, they also have significantly lower odds of initiating LTBI treatment, which could be due to costs associated with it. Although the actual treatment regime is offered to all LTBI cases and provided without payment, foreign nationals are required to pay for follow-up laboratory tests and consultations as part of the treatment monitoring process. These costs, and that LTBI treatment is not mandatory, makes it less likely for foreign nationals to comply. Although it is not legally required, there are cases where certain employers chose to terminate employment contracts for foreign workers diagnosed with LTBI. In Brunei, such termination is required only for those diagnosed with active TB disease [23, 24]. As contacts with LTBI diagnosis were not actively followed-up after being offered LTBI treatment, its extent towards treatment initiation is unclear.

It should be noted that foreign workers are routinely screened with chest X-ray only to identify TB lesions upon their entry into Brunei. These workers tend to come from high TB-burden countries; foreign nationals constituted 28.0% of Brunei’s work force, with a significant proportion employed as low-skilled workers in construction, wholesale and retail trade [25]. As TST is unable to distinguish between recent or past infection, it is probable that LTBI detected during contact investigations may not be attributed to the particular index case. However, history of Bacille Calmette-Guérin (BCG) vaccination for both locals and foreign nationals are unlikely to differ as Brunei (like many parts of the world) routinely provides BCG vaccination at birth, with population coverage > 95% since the early 2000s [5]. Thus, history of vaccine uptake should not be a factor to withhold TST [3]. Providing LTBI screening and treatment for foreign workers at entry would be important step towards TB elimination [26].

In addition, we found that the adjusted odds of being diagnosed with LTBI was increased by 2-3 times if the contact resided in Belait and Tutong districts, when compared to Brunei-Muara. This finding could be explained by operational challenges that is prevalent in the latter district. Brunei-Muara is the most populated district where 69.7% of the country’s population reside [4]. Also, TB health officers/doctors from Brunei-Muara are also responsible to man the DOTS clinic at Temburong district. Such issues were minimal when compared to both Belait and Tutong districts with smaller catchment populations.. Hence, human resource constraints to keep hardcopy records of certain contact investigations were possible. Also, the DOTS clinic at Brunei-Muara was the only clinic affected with relocation twice during the study period; relevant hardcopy records could be lost during these relocations. Digitalising future records from contact investigations into a database is one feasible and effective way to address this issue.

Interestingly, we observed that index factors play a more significant role with regards to LTBI treatment completion. LTBI cases detected from younger index cases (< 15 years old) had lower adjusted odds of completing LTBI treatment. One possible explanation could be the lower perceived severity as presenting symptoms of younger PTB cases tend to be less severe when compared to adults [27]. Contacts of younger PTB cases may assume that it is not necessary to complete LTBI treatment based on the assumption that their index case was not very infectious. However, it should be noted that there are only 9 index cases of ages <15 years, indicating that this result should be treated with caution. Alternate versions of the same model were analysed (collapsing age-groups and treating age as numeric variable) but we decided to use this version due to its lower QICu value. In a similar manner, perceived severity of the disease (again possibly based on presenting symptoms and/or initial stay at an isolation centre of the index case) could possibly explain significantly high odds of completing LTBI treatment for contacts of smear positive PTB index cases. Patients were not generally informed about their smear positive status (unless inquired by the patient). Also, higher adjusted odds of LTBI treatment completion for contacts of local PTB index could indicate high compliance due to low barriers for treatment completion; Presumably majority of the contacts of local PTB index cases are locals themselves, and are therefore eligible for treatment and follow-up laboratory tests at no monetary cost. Lastly, higher LTBI treatment completion rates for the latter half of the study period (2014-2018) is mainly due to the strengthening of the whole programme after the national TB guideline revision in 2013, which has led to a more systematic and dedicated approach towards LTBI patient counselling and follow-up.

In our study, 5 LTBI patients (0.5%) progressed to active TB, a proportion similar to a national database-linkage study in Taiwan [28]. However, we believe that this progression may not be entirely attributable to TB reactivation, as all except 1 have completed their full course of LTBI treatment and were otherwise healthy individuals with no co-morbidities. Also, 2/5 patients developed active TB disease within the first 2 years of LTBI, the time period with the highest risk of reactivation [29]. Rather, subsequent re-infection could be more probable. Household members of 2 LTBI patients had prior history of PTB infection after they themselves have completed LTBI treatment. These repeated significant exposures within the household may possibly lead to re-infection. Further studies that include molecular typing of *Mycobacterium Tuberculosis* strains will be needed to investigate this hypothesis.

This study has a number of limitations. First, contact tracing data was retrospectively extracted from hardcopy records and manually entered into an excel sheet. Hence, data entry errors were possible. Efforts were made to minimise this, including initial checks among data entry staff to ensure completeness, and further checks to clarify and remove duplicate entries. Second, we could not account for (and also unable to assess the impact of) possible missing information resulting from either the lack of data recording or loss of hardcopy records during relocation. Third, the nature of the dataset (where there is a separate set of cases and contacts) makes it difficult to evaluate the yield of active TB cases from contact investigations. This percentage is likely to be very low based on field experience, similar to reports from other countries [2]. Fourth, as LTBI diagnosis in this study relied solely on TST screening, the extent of this screening on routine contact investigations could be limited during periods of PPD supply shortage. Such shortage occurred in Brunei between 2015 and 2018, and also at a global level [3, 30] and has led to restrictions on LTBI testing for only contacts of smear positive PTB and child TB cases. Despite these restrictions, the proportion of LTBI diagnosis across settings remain quite constant and consistent when compared to 2009-13, except for settings other than household, workplaces and schools. Interferon-γ release assays (IGRAs) could be considered as an alternative during periods of global PPD supply disruption. Either tests are recommended for LTBI testing by the WHO [3], although both tests have their own imperfections and have limited evidence for the predicting progression to active TB disease [31]. Lastly, our findings on the proportion of LTBI cases who progressed to active TB disease may not accurately reflect the true number, as the matching process was performed manually and some cases may have been missed or not taken into account. Establishing a centralized national TB database will be a helpful platform to conduct similar studies in the future.

In conclusion, we found that 9.9% (n= 1047) of all contacts of PTB index cases identified during the 10-year study period were diagnosed as LTBI, out of which only 43.0% (n= 450) initiated LTBI treatment. Among those who initiated, 74.0% of them completed LTBI treatment. LTBI burden is disproportionately high towards foreign nationals, with higher odds of LTBI diagnosis but lower odds of LTBI treatment initiation. Determining the reasons of not initiating LTBI treatment would help the programme to strategize ways to improve LTBI treatment uptake. If these reasons are related to monetary costs, discussions to include LTBI treatment as part of foreign workers’ insurance coverage could possibly be initiated. Also, starting a digital linkage database between cases and contacts will be helpful to better evaluate the number of contacts who progressed towards active TB disease, and also to monitor the impact of contact investigation in the local context. Molecular typing would be useful to determine the risk of LTBI reactivation versus reinfection among LTBI cases, particularly within enclosed settings.

## Data Availability

All data produced in the present study are available upon reasonable request to the corresponding author.

## Acknowledgment

The authors would like to thank Madam Hardew Kaur, Nur Syazwani Mohd Bahrin, as well as all staff at NTCC and DOTS clinics for their assistance during data collection. We are also grateful for our research assistants (Sa’adatul Humairah Damit, Nurul Aqilah Md Zain, and Nurazirah Abdul Azis) who digitally compiled and checked the datasets used in this study.

## Author contributions

LC, RAH and KT conceived the study.

RAH, KSK and KT collected the data.

LC and analysed the data and drafted the manuscript.

All authors read, revised and approved the manuscript.

## Patient and Public Involvement

Patients or the public were not involved in the design, or conduct, or reporting, or dissemination plans of our research.

